# Ischemic stroke after bivalent mRNA COVID-19 vaccination and influenza vaccination during the 2022-2023 season: a multi-site self-controlled case series study

**DOI:** 10.64898/2026.05.20.26353716

**Authors:** Stanley Xu, Lina S. Sy, Vennis Hong, Paddy Farrington, Sungching C. Glenn, Sunhea Kim, Denison S. Ryan, Julia E Tubert, Paulina Tong, Bruno J. Lewin, Hung Fu Tseng, Alexandra Carbayo, Cameron Davis, Navdeep S. Sangha, Edward A Belongia, Maria E. Sundaram, Jennifer C. Nelson, Matthew F. Daley, Nicola P. Klein, Bruce Fireman, Jacob Haapala, Laura P. Hurley, Stephanie A. Irving, Noelle M. Cocoros, Eric S. Weintraub, Jonathan Duffy, Lei Qian

## Abstract

**Background:** The Vaccine Safety Datalink (VSD) detected a statistical signal for ischemic events (ischemic stroke or transient ischemic attack) following bivalent mRNA COVID-19 vaccination through prospective surveillance during 2022-2023. Although multiple studies from other surveillance systems and countries reported no increased risk, important methodological limitations remained. This U.S. study addressed those limitations by evaluating the ischemic stroke risk following bivalent mRNA COVID-19 vaccination, influenza vaccination, and their same-day coadministration using event-dependent self-controlled case series (SCCS) design.

**Methods:** Study outcomes included first-ever ischemic stroke (primary outcome), first-in-1-year ischemic stroke (secondary outcome), and ischemic events (exploratory outcomes), identified using ICD-10-CM codes in inpatient and emergency department settings during September 1, 2022–March 31, 2023, among individuals aged ≥12 years across eight VSD sites. Analyses were conducted separately for Pfizer-BioNTech and Moderna bivalent vaccines, with relative incidences (RI) and 95% confidence intervals (CI) estimated for 1–21-day and 1–42-day risk intervals, using person-time outside these intervals as the control period. Subgroup analyses were performed by age group (12–64, >65 years) and history of documented SARS-CoV-2 infection.

**Results:** A total of 6,510 first-ever ischemic strokes were identified among more than 6.8 million participants. Among recipients of Pfizer-BioNTech bivalent COVID-19 and influenza vaccines, no statistically significant increased risk of first-ever ischemic stroke was observed following bivalent COVID-19 vaccination (RI = 0.94; 95% CI: 0.63–1.41), influenza vaccination (RI = 0.95; 95% CI: 0.82–1.10), or same-day coadministration (RI = 1.15; 95% CI: 0.88–1.49) within 1– 21-day risk intervals; findings were similar for 1–42-day intervals. Comparable null results were observed for Moderna vaccines and across all subgroups, secondary, and exploratory outcomes.

**Conclusion:** No increased risk of ischemic stroke was found following bivalent mRNA COVID-19 vaccination, influenza vaccination, or their coadministration in this multi-site SCCS study. These findings are consistent with previous studies and underscore the importance of continued vaccine safety monitoring.

**Highlights:**

- We evaluated ischemic stroke risk after bivalent mRNA COVID-19 vaccination, influenza vaccination, and their coadministration.
- No increased risk of ischemic stroke was observed within 1–21 or 1–42 days following these vaccination exposures.
- Findings were consistent across vaccine type, age group, and prior SARS-CoV-2 infection status.
- These results support the safety of bivalent mRNA COVID-19 vaccines with respect to ischemic stroke.

## 1. BACKGROUND

The U.S. Food and Drug Administration (FDA) authorized the use of bivalent mRNA COVID-19 vaccines in late August 2022, introducing updated formulations from Pfizer-BioNTech for individuals aged 12 years and older, and from Moderna for those aged 18 years and older [1,2]. The bivalent vaccine was initially recommended as a single booster dose for people who had completed a primary series or received a prior monovalent booster, typically at least 2 months earlier [2]. The rollout occurred during a period of waning immunity from earlier vaccinations, and the updated vaccines were expected to restore and enhance protection against circulating SARS-CoV-2 variants [3-5].

In the United States, the Vaccine Safety Datalink (VSD) has monitored COVID-19 vaccine safety since vaccinations began in December 2020. In late 2022, VSD’s rapid cycle analyses (RCA), which used a concurrent comparator design [6], preliminarily detected a statistical signal for ischemic events (defined as ischemic stroke [i.e., cerebral infarction] or transient ischemic attack [TIA]) following the Pfizer-BioNTech COVID-19 bivalent booster among adults aged 65 years and older. Signal evaluation analyses suggested a potential higher risk when bivalent COVID-19 vaccines were coadministered with a high-dose or adjuvanted influenza vaccine [7]. This signal was not considered a confirmed safety concern, in part because the underlying events had not been chart-validated and residual confounding was not fully addressed. As a result, it prompted further investigation, including several follow-up studies summarized below as well as the current study.

Several other studies did not identify an increased risk of ischemic stroke following COVID-19 bivalent vaccination. Surveillance using V-safe and the Vaccine Adverse Event Reporting System (VAERS) found that the safety profile of bivalent mRNA COVID-19 vaccines in individuals aged 12 years and older was similar to that of monovalent COVID-19 booster vaccinations [8]. A TreeScan data mining analysis in the VSD among enrollees aged ≥5 years who received Pfizer-BioNTech or Moderna bivalent COVID-19 vaccines examined a broad range of adverse events and identified no increased risk of ischemic events [9]. A U.S. cohort study using TriNetX electronic health record data of adults aged ≥65 years reported that, within both 1–21 and 22–42 days after vaccination, Pfizer-BioNTech bivalent recipients had a lower risk of ischemic stroke compared with recipients of Pfizer-BioNTech or Moderna monovalent boosters, with no significant difference between the Pfizer and Moderna bivalent groups [10]. A French cohort study similarly found that bivalent vaccination was not associated with increased risk of ischemic stroke, hemorrhagic stroke, myocardial infarction, or pulmonary embolism when compared with monovalent booster vaccination [11].

The self-controlled case series (SCCS) method [12-14] was also used in several studies to assess the risk of ischemic events after COVID-19 bivalent vaccination while implicitly adjusting for time-invariant confounders. An SCCS study in England found no indication of increased risk of ischemic events, including ischemic stroke or TIA, within 21 days of receipt of either mRNA bivalent vaccine [15]. Similarly, an SCCS study in Israel detected no increased risk of ischemic stroke following monovalent or bivalent Pfizer-BioNTech booster doses [16].

In contrast, a smaller number of studies have reported positive associations or suggestive trends. Among Medicare beneficiaries aged ≥65 years who received both a COVID-19 bivalent vaccine and a high-dose or adjuvanted influenza vaccine, an elevated risk of nonhemorrhagic stroke (i.e., ischemic stroke) was observed after Pfizer-BioNTech bivalent vaccination, within a 22–42 days risk interval compared with the 43–90-day control interval [17]. A single-site SCCS study did not find a statistically significant increased risk of ischemic events, but the point estimates were elevated within the 1–42-day risk interval among individuals aged 12–64 years who received same-day coadministration of Pfizer-BioNTech bivalent and influenza vaccines and among Moderna bivalent recipients aged 12–64 years with prior SARS-CoV-2 infection, although these associations did not reach statistical significance [18].

However, previous studies had important limitations. Some did not examine ischemic stroke as a distinct outcome but instead evaluated composite ischemic events (ischemic stroke or TIA) [9, 15,18] and others did not have sufficient sample size [18] to draw definitive conclusions. Additionally, most studies did not account for a history of SARS-CoV-2 infection or assess the risk of ischemic events following coadministration of influenza vaccine and COVID-19 vaccines.

This multi-site study aimed to assess the risk of ischemic stroke and TIA following bivalent COVID-19 vaccination alone, influenza vaccination alone, and coadministration of the two vaccines among individuals enrolled in eight VSD sites using an event-dependent SCCS design. This signal-evaluation study differs from the RCA concurrent comparator approach because the SCCS method implicitly adjusts for time-invariant confounding. We hypothesized that bivalent mRNA COVID-19 vaccination, influenza vaccination, and their same-day coadministration are not associated with an increased risk of ischemic stroke or TIA during predefined post-vaccination risk intervals compared with control intervals within individuals.

## 2. METHODS

### 2.1. Study Population and Study Period

This study utilized electronic health records from a cohort of individuals aged ≥12 years enrolled at eight VSD sites: Denver Health, HealthPartners (Minnesota), Kaiser Permanente (KP) Colorado, KP Northern California, KP Northwest, KP Southern California, KP Washington, and Marshfield Clinic (Wisconsin). From the cohort, the SCCS analytic datasets were created including only individuals who experienced ischemic events during September 1, 2022–March 31, 2023. To be eligible, individuals must have completed a COVID-19 vaccine primary series and received their last monovalent dose at least 60 days prior to September 1, 2022. Additionally, individuals were required to have at least one year of continuous enrollment (allowing for a 31-day gap) prior to September 1, 2022, except at Denver Health, where individuals were required to have at least one primary care visit in the prior 18 months. We excluded individuals who received influenza vaccination in the 2 weeks prior to September 1, 2022, to minimize the impact of influenza vaccination before the observation period on risk estimation; receipt of vaccines other than influenza or COVID-19 vaccines during this period was permitted.

This study was reviewed and approved by the institutional review boards of all participating sites and was conducted consistent with federal law and CDC policy (See 45C.F.R. part 46.102(l)(2), 21C.F.R. part 56).

### 2.2. Exposure and Observation Period

The exposures were the administration of the Pfizer-BioNTech or Moderna bivalent COVID-19 vaccines, as well as any influenza vaccine, during the period from September 1, 2022 through March 31, 2023. For individuals who received both vaccines during the observation period, we identified instances of coadministration—either on the same day or in close proximity (i.e., coadministration on different days but within overlapping risk intervals)—and excluded those with coadministration in close proximity for the reasons described in the Statistical Analyses section. The observation period for recipients of bivalent COVID-19 vaccines and influenza vaccine started on September 1, 2022, and ended on March 31, 2023, or upon death, receipt of a second bivalent or second influenza vaccine dose, or disenrollment, whichever occurred first.

To adjust for seasonality effects, we also included ischemic events among eligible individuals who did not receive a bivalent COVID-19 vaccine or influenza vaccine during the study period (non-vaccinees). For non-vaccinees, the observation period started on September 1, 2022, and ended on March 31, 2023, or upon death or disenrollment, whichever occurred first.

### 2.3. Outcome Definition

We identified ischemic events during the observation period (index events), defined as ischemic stroke or TIA, in emergency department or inpatient settings using International Classification of Diseases, Tenth Revision (ICD-10) codes: G45.8 and G45.9 for TIA, and I63* for ischemic stroke. We excluded ischemic events attributed to other possible causes documented in electronic health records, including those occurring within 30 days of SARS-CoV-2 infection (confirmed by a positive laboratory test or a COVID-19 diagnosis), given well-documented evidence of substantially elevated ischemic stroke risk in the short-term period following SARS-CoV-2 infection [19,20], to reduce potential confounding by infection-related ischemic risk and avoid biasing vaccine safety estimates. We also adjusted the automated onset dates if certain diagnoses—such as status post administration of tissue plasminogen activator in a different facility, headache, speech disturbances, facial weakness, weakness, dizziness and giddiness, altered mental status, transient alteration of awareness, hemiplegia, unspecified visual disturbance, or sudden visual loss— were present the day prior to the event [6,18].

The primary outcome was first-ever ischemic stroke, defined as the first ischemic stroke recorded for an individual with no documented ischemic stroke prior to the index event in the ICD-10-CM era (i.e., after October 2016). This outcome was chosen to reflect more severe disease and minimize potential outcome misclassification due to inadvertent inclusion of TIA and avoid capture of pre-existing conditions. The secondary outcome was first-in-1-year ischemic stroke, defined as an ischemic stroke occurring in an individual with no documented ischemic stroke in the one year prior to the index event. This secondary outcome captured ischemic stroke events among individuals with prior ischemic stroke for more than 1 year and addressed the potential increased risk in a population predisposed to ischemic stroke. Exploratory outcomes included composite ischemic events, defined as first-ever ischemic stroke or TIA, and first-in-1-year ischemic stroke or TIA (Figure 1).

**Figure 1.**
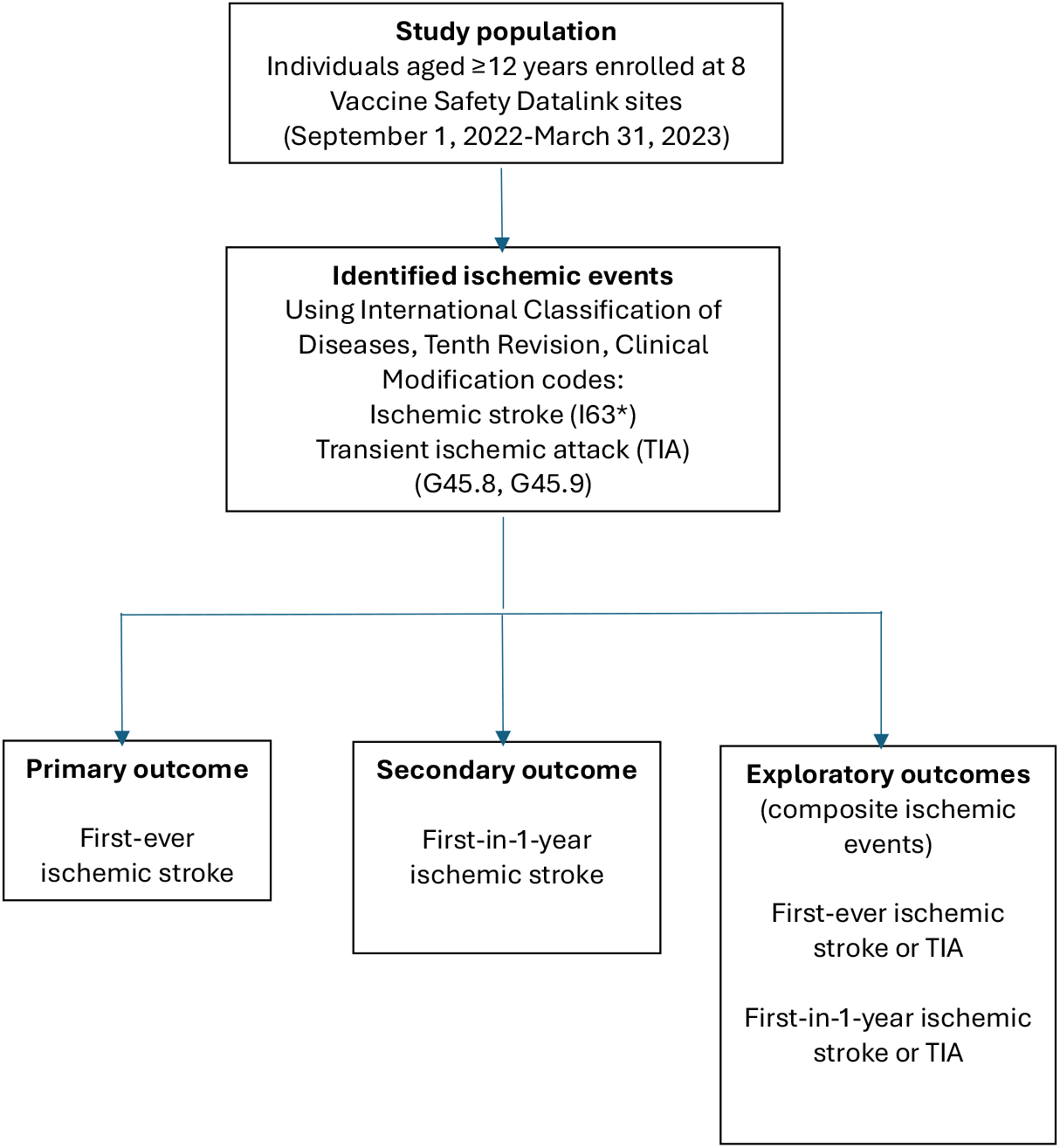
Identification of ischemic events and definition of study outcomes

### 2.4. Statistical Analyses

Demographic characteristics of individuals with ischemic stroke events during the study period were described separately for recipients of Pfizer-BioNTech bivalent COVID-19 vaccine alone (defined as receipt of a COVID-19 vaccine without influenza vaccination, but allowing receipt of other vaccines), recipients of Moderna bivalent COVID-19 vaccine alone (similarly defined), recipients of influenza vaccine alone (without COVID-19 vaccination, but allowing other vaccines), recipients of Pfizer -BioNTech bivalent COVID-19 vaccine and influenza vaccine (also allowing other vaccines), recipients of Moderna bivalent COVID-19 vaccine and influenza vaccine (also allowing other vaccines), and non-vaccinees of bivalent COVID-19 and influenza vaccines (allowing receipt of other vaccines). Given that prior study outcomes might influence the probability of future vaccination, we employed an event-dependent SCCS approach tailored for event-dependent exposures [21-23]. This event-dependent SCCS used a pseudo-likelihood approach within a counterfactual framework to estimate relative incidence (RI) and 95% confidence intervals (CI), comparing the risk intervals to the corresponding control intervals within individuals.

Coadministration of COVID-19 vaccine and influenza vaccine posed challenges in the event-dependent SCCS analysis, particularly coadministration not on the same day but in close proximity with overlapping risk intervals. In this SCCS study, we treated each vaccination as a distinct exposure to assess risk of study outcomes after bivalent COVID-19 vaccination alone, influenza vaccination alone, and same-day coadministration in a single event-dependent SCCS analysis [22]. However, the event-dependent SCCS method as currently available could not accommodate cases of coadministration in close proximity with overlapping risk intervals. Therefore, cases of coadministration in close proximity with overlapping risk intervals but not on the same day were excluded from further analyses.

The risk intervals were pre-specified as 1–21 days and 1–42 days after bivalent COVID-19 vaccination alone, influenza vaccination alone, and same-day coadministration in an event-dependent SCCS analysis, with person-time outside of these risk intervals, including pre-vaccination person-time and post-risk person-time, serving as the control interval. We illustrated the risk and control intervals for seven vaccination scenarios in Figure 2. We excluded Day 0 (vaccination date) from the risk and control intervals because without conducting chart review, it would not be possible to determine whether the vaccination or the study outcomes occurred first on Day 0. To maintain consistency, we excluded Day 0 from the risk and control intervals for bivalent COVID-19 vaccination, influenza vaccination, and their same-day coadministration. In these analyses, study outcomes occurring among eligible non-vaccinees were included to adjust for seasonality by incorporating calendar month into the models. Age was not adjusted as a time-varying covariate given the relatively short observation period of 7 months.

**Figure 2.**
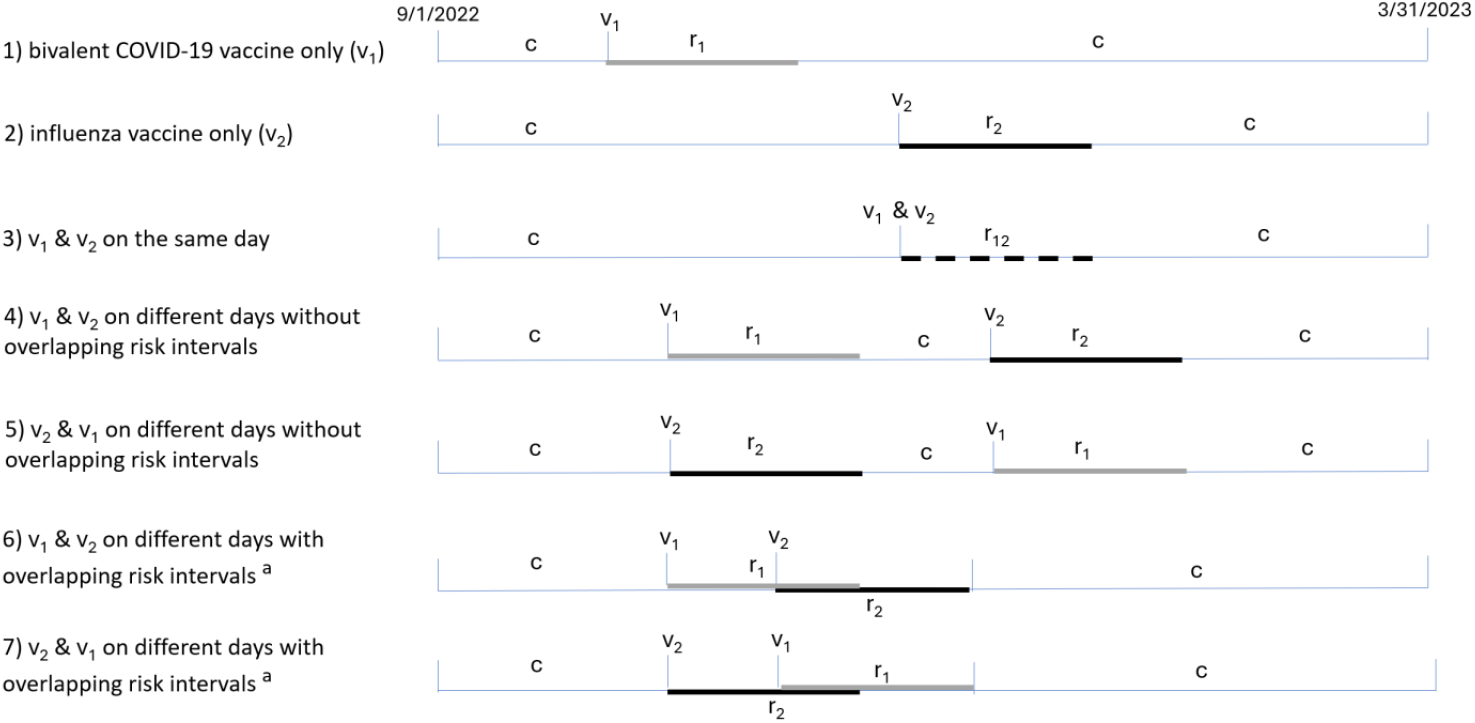
Risk and control intervals for seven scenarios among individual who received bivalent COVID-19 and/or influenza vaccines: 1) an individual who received a bivalent COVID-19 vaccine (v_1_) without influenza vaccination with a risk interval r_1_, 2) an individual who received a influenza vaccine (v_2_) without a bivalent COVID-19 vaccine with a risk interval r_2_, 3) an individual who received these two vaccines on the same day with a risk interval r_12_, 4) and 5) individuals who received these two vaccines on different days without overlapping risk intervals, and 6) and 7) individuals who received these two vaccines on different days with overlapping risk intervals. The person-time outside of the risk intervals serves as the control interval (c). ^a^ Cases were excluded in self-controlled case series analyses if coadministration of the two vaccines was in close proximity (but not same day) with overlapping risk intervals.

Subgroup analyses were performed by age group (12-64 years and ≥65 years) because VSD’s concurrent-comparator RCA detected a statistical signal among adults aged ≥65 years, and by history of documented SARS-CoV-2 infection (yes/no) within one year prior to September 1, 2022. Documented SARS-CoV-2 infection was defined as a positive laboratory test or a COVID-19 diagnosis in the medical record; results from home COVID-19 tests were not included.

Chart reviews of study outcomes were planned to enable analyses including only confirmed cases if a significantly elevated RI was observed in analyses of electronically identified events.

Analyses were conducted using SAS version 9.4 (SAS Institute), and the event-dependent SCCS models were fitted using the R package (R Core Team) SCCS [24].

This study was reviewed and approved by the institutional review boards of all participating sites consistent with federal law and CDC policy (See 45C.F.R. part 46.102(l)(2), 21C.F.R. part 56). A waiver of informed consent was granted due to the study posing minimal risk to participants.

## 3. RESULTS

### 3.1. Characteristics of those who had first-ever ischemic stroke

Among more than 6.8 million individuals, 6,510 experienced first-ever ischemic stroke during September 1, 2022–March 31, 2023. Of these cases, 182 (2.8%) received only Pfizer-BioNTech bivalent COVID-19 vaccine, 78 (1.2%) received only Moderna bivalent COVID-19 vaccine, 2,123 (32.6%) received only the influenza vaccine, 1,704 (26.2%) received both the Pfizer-BioNTech bivalent COVID-19 vaccine and the influenza vaccine, 863 (13.3%) received both the Moderna bivalent COVID-19 vaccine and the influenza vaccine, and 1,560 (24.0%) did not receive any of these vaccines (Table 1).

**Table 1.** Characteristics of individuals aged 12 years and older who had first-ever ischemic stroke across 8 Vaccine Safety Datalink sites during September 1, 2022–March 31, 2023.^1^.

|  | Recipients of only Pfizer -BioNTech bivalent COVID-19 vaccine, no. (%) | Recipients of only Moderna bivalent COVID-19 vaccine, no. (%) | Recipients of only influenza vaccine, no. (%) | Recipients of Pfizer - BioNTech bivalent COVID-19 vaccine and influenza vaccine<sup>2</sup>, no. (%) | Recipients of Moderna bivalent COVID-19 vaccine and influenza vaccine<sup>2</sup>, no. (%) | Non-vaccinees<sup>3</sup>, no. (%) |
| --- | --- | --- | --- | --- | --- | --- |
| Overall | 182 | 78 | 2,123 | 1,704 | 863 | 1,560 |
| Age (in years) |  |  |  |  |  |  |
| 12-17 | 0 (0) | 0 (0) | 5 (0) | 3 (0) | 0 (0) | 3 (0) |
| 18-44 | 8 (4) | 2 (3) | 140 (7) | 49 (3) | 19 (2) | 179 (11) |
| 45-64 | 62 (34) | 27 (35) | 627 (30) | 357 (21) | 187 (22) | 607 (39) |
| 65-74 | 55 (30) | 24 (31) | 517 (24) | 497 (29) | 254 (29) | 328 (21) |
| 75+ | 57 (31) | 25 (32) | 834 (39) | 798 (47) | 403 (47) | 443 (28) |
| Sex |  |  |  |  |  |  |
| Female | 92 (51) | 40 (51) | 1117 (53) | 883 (52) | 432 (50) | 793 (51) |
| Male | 90 (49) | 38 (49) | 1006 (47) | 821 (48) | 431 (50) | 767 (49) |
| Race/ethnicity |  |  |  |  |  |  |
| Hispanic | 30 (16) | 14 (18) | 579 (27) | 263 (15) | 175 (20) | 345 (22) |
| Non-Hispanic White | 83 (46) | 31 (40) | 944 (44) | 1013 (59) | 457 (53) | 684 (44) |
| Non-Hispanic Asian | 24 (13) | 12 (15) | 252 (12) | 209 (12) | 97 (11) | 157 (10) |
| Non-Hispanic Black | 28 (15) | 14 (18) | 213 (10) | 126 (7) | 85 (10) | 243 (16) |
| Missing | 8 (4) | 2 (3) | 30 (1) | 19 (1) | 20 (2) | 52 (3) |
| Multiple/Other | 9 (5) | 5 (6) | 105 (5) | 74 (4) | 29 (3) | 79 (5) |

<sup>1</sup> Vaccination status was assessed at the end of follow-up, regardless of when the event occurred, and Day 0 cases were included in Table 1; <sup>2</sup>Those who received bivalent COVID-19 vaccine and influenza vaccine on the same day or on different days during September 1, 2022–March 31, 2023, regardless of the interval between vaccinations; <sup>3</sup>Those who did not receive a bivalent vaccine or influenza vaccine during September 1, 2022–March 31, 2023.

Tables S1 and S2 summarize the total number of vaccinated cases, the number of Day 0 cases, the number of cases with coadministration in close proximity with overlapping risk intervals, as well as the final case counts included in SCCS analyses. For example, among 4,009 vaccinated first-ever ischemic stroke events, 89 events occurred on Day 0 and 452 involved coadministration of Pfizer-BioNTech bivalent COVID-19 and influenza vaccines within 1–21 days apart, leaving 3,468 cases eligible for the 1–21-day SCCS analysis. When applying a 1– 42-day interval, 751 cases involved non-same-day coadministration in close proximity with overlapping risk intervals and were excluded, resulting in 3,169 cases included in the SCCS analysis.

### 3.2. Primary outcome first-ever ischemic stroke

For the recipients of Pfizer-BioNTech bivalent vaccination and influenza vaccination, with 3,468 first-ever ischemic strokes among vaccinees, the overall (all ages pooled) analysis showed non-statistically significant RIs of 0.94 (95% CI, 0.63–1.41), 0.95 (95% CI, 0.82–1.10), and 1.15 (95% CI, 0.88–1.49) within 1–21 days after Pfizer-BioNTech bivalent vaccination, influenza vaccination, and same-day coadministration, respectively (Table 2). Similarly, no elevated risk was identified for the 1–42 day risk interval or in subgroup analyses. We also presented age-stratified estimates (12–64 and ≥65 years) because baseline ischemic stroke risk is age-dependent and the original VSD signal was detected among adults aged ≥65 years; pooling across age groups could mask age-specific patterns. Although the point estimates of RI tended to be higher in some subgroups (e.g., those with prior SARS-CoV-2 infection or same-day coadministration), they were not statistically significant.

**Table 2.** Relative incidences of electronically identified first-ever ischemic stroke in the 21 days and 42 days after Pfizer-BioNTech bivalent COVID-19 vaccination and influenza vaccination among members of 8 Vaccine Safety Datalink sites during September 1, 2022–March 31, 2023.

|  |  | Relative incidence for ≥12 years old |  |  | Relative incidence for 12-64 years old |  |  | Relative incidence for ≥65 years old |  |  |
| --- | --- | --- | --- | --- | --- | --- | --- | --- | --- | --- |
| Risk interval | Analyses | After COVID-19 vaccination | After influenza vaccination | After same-day coadministration | After COVID-19 vaccination | After influenza vaccination | After same-day coadministration | After COVID-19 vaccination | After influenza vaccination | After same-day coadministration |
| 1-21 days | Overall | 0.94 (0.63–1.41) | 0.95 (0.82–1.10) | 1.15 (0.88–1.49) | 1.04 (0.52–2.10) | 1.08 (0.84–1.39) | 0.89 (0.51–1.56) | 0.90 (0.55–1.48) | 0.91 (0.76–1.09) | 1.26 (0.94–1.70) |
|  | With prior SARS-CoV-2 infection<sup>1</sup> | 1.07 (0.36–3.18) | 1.06 (0.75–1.50) | 1.46 (0.76–2.78) | 2.98 (0.71–12.53) | 1.38 (0.85–2.25) | 1.18 (0.36–3.83) | 0.37 (0.05–2.95) | 0.87 (0.53–1.44) | 1.59 (0.73–3.48) |
|  | Without prior SARS-CoV-2 infection | 0.93 (0.60–1.44) | 0.93 (0.79–1.09) | 1.11 (0.83–1.47) | 0.79 (0.34–1.86) | 1.00 (0.75–1.34) | 0.83 (0.44–1.56) | 0.99 (0.59–1.66) | 0.92 (0.76–1.11) | 1.22 (0.88–1.69) |
| 1-42 days | Overall | 0.82 (0.56–1.20) | 0.92 (0.81–1.04) | 1.04 (0.84–1.30) | 0.93 (0.51–1.72) | 1.09 (0.88–1.35) | 1.02 (0.67–1.55) | 0.77 (0.48–1.24) | 0.85 (0.73–1.00) | 1.06 (0.82–1.37) |
|  | With prior SARS-CoV-2 infection<sup>1</sup> | 1.57 (0.65–3.81) | 1.08 (0.80–1.46) | 1.51 (0.87–2.62) | 3.16 (0.78–12.82) | 1.17 (0.74–1.84) | 2.11 (0.82–5.47) | 1.05 (0.32–3.50) | 1.04 (0.70–1.56) | 1.27 (0.64–2.52) |
|  | Without prior SARS-CoV-2 infection | 0.74 (0.48–1.12) | 0.89 (0.77–1.02) | 0.99 (0.78–1.25) | 0.72 (0.35–1.50) | 1.07 (0.84–1.36) | 0.86 (0.54–1.38) | 0.74 (0.44–1.24) | 0.83 (0.70–0.97) | 1.03 (0.79–1.36) |

<sup>1</sup> Had SARS-CoV-2 infection (i.e., SARS-CoV-2 positive laboratory test or a COVID-19 diagnosis) during the year prior (September 1, 2021–March 31, 2022).

Table 3 shows the results for the Moderna bivalent vaccination and influenza vaccination, also indicating no elevated risk for first-ever ischemic stroke following either vaccination alone or same-day coadministration.

**Table 3.**
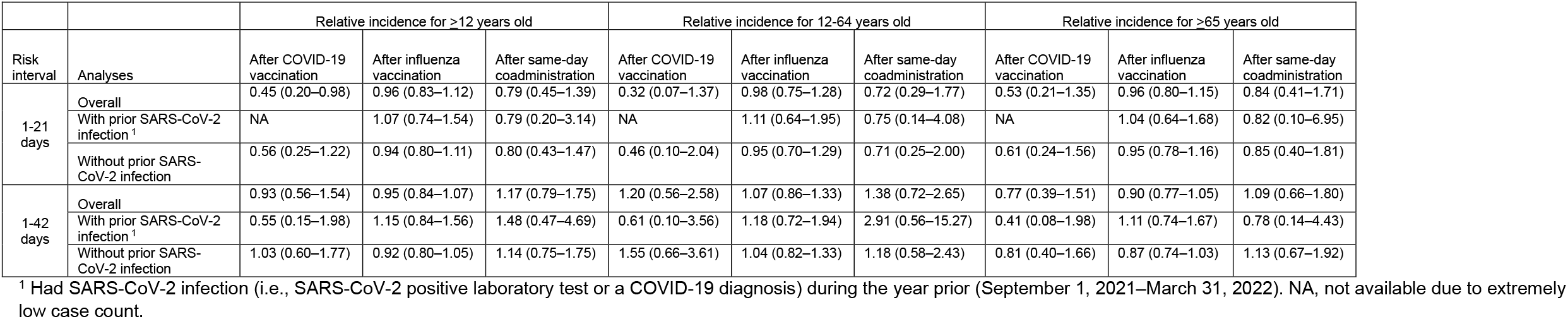
Relative incidences of electronically identified first-ever ischemic stroke in the 21 days and 42 days after Moderna bivalent COVID-19 vaccination and influenza vaccination among members of 8 Vaccine Safety Datalink sites during September 1, 2022–March 31, 2023.

|  |  | Relative incidence for ≥12 years old |  |  | Relative incidence for 12-64 years old |  |  | Relative incidence for ≥65 years old |  |  |
| --- | --- | --- | --- | --- | --- | --- | --- | --- | --- | --- |
| Risk interval | Analyses | After COVID-19 vaccination | After influenza vaccination | After same-day coadministration | After COVID-19 vaccination | After influenza vaccination | After same-day coadministration | After COVID-19 vaccination | After influenza vaccination | After same-day coadministration |
| 1-21 days | Overall | 0.45 (0.20–0.98) | 0.96 (0.83–1.12) | 0.79 (0.45–1.39) | 0.32 (0.07–1.37) | 0.98 (0.75–1.28) | 0.72 (0.29–1.77) | 0.53 (0.21–1.35) | 0.96 (0.80–1.15) | 0.84 (0.41–1.71) |
|  | With prior SARS-CoV-2 infection<sup>1</sup> | NA | 1.07 (0.74–1.54) | 0.79 (0.20–3.14) | NA | 1.11 (0.64–1.95) | 0.75 (0.14–4.08) | NA | 1.04 (0.64–1.68) | 0.82 (0.10–6.95) |
|  | Without prior SARS-CoV-2 infection | 0.56 (0.25–1.22) | 0.94 (0.80–1.11) | 0.80 (0.43–1.47) | 0.46 (0.10–2.04) | 0.95 (0.70–1.29) | 0.71 (0.25–2.00) | 0.61 (0.24–1.56) | 0.95 (0.78–1.16) | 0.85 (0.40–1.81) |
| 1-42 days | Overall | 0.93 (0.56–1.54) | 0.95 (0.84–1.07) | 1.17 (0.79–1.75) | 1.20 (0.56–2.58) | 1.07 (0.86–1.33) | 1.38 (0.72–2.65) | 0.77 (0.39–1.51) | 0.90 (0.77–1.05) | 1.09 (0.66–1.80) |
|  | With prior SARS-CoV-2 infection<sup>1</sup> | 0.55 (0.15–1.98) | 1.15 (0.84–1.56) | 1.48 (0.47–4.69) | 0.61 (0.10–3.56) | 1.18 (0.72–1.94) | 2.91 (0.56–15.27) | 0.41 (0.08–1.98) | 1.11 (0.74–1.67) | 0.78 (0.14–4.43) |
|  | Without prior SARS-CoV-2 infection | 1.03 (0.60–1.77) | 0.92 (0.80–1.05) | 1.14 (0.75–1.75) | 1.55 (0.66–3.61) | 1.04 (0.82–1.33) | 1.18 (0.58–2.43) | 0.81 (0.40–1.66) | 0.87 (0.74–1.03) | 1.13 (0.67–1.92) |

<sup>1</sup> Had SARS-CoV-2 infection (i.e., SARS-CoV-2 positive laboratory test or a COVID-19 diagnosis) during the year prior (September 1, 2021–March 31, 2022). NA, not available due to extremely low case count.

### 3.3. Secondary outcome first-in-1-year ischemic stroke

There were 3,847 first-in-1-year ischemic strokes among vaccinees, 379 more than first-ever ischemic strokes. For both Pfizer-BioNTech bivalent vaccination and influenza vaccination, we found no evidence of increased risk for first-in-1-year ischemic stroke following either vaccine alone or same-day coadministration. This held true in overall analyses and in stratified analyses by age group (12– 64 years old and ≥65 years old) and prior SARS-CoV-2 infection status, for both 1–21-day and 1–42-day risk intervals (Table 4). Elevated point estimates were observed within 42 days after Pfizer-BioNTech bivalent vaccination (RI=3.28 [95%CI, 0.81–13.30]) and same-day coadministration (RI=2.09 [95% CI, 0.85–5.14]) in individuals aged 12-64 years with prior SARS-CoV-2 infection, although the results were not statistically significant and the confidence intervals were wide.

**Table 4.** Relative incidences of electronically identified first-in-1-year ischemic stroke in the 21 days and 42 days after Pfizer-BioNTech bivalent COVID-19 vaccination and influenza vaccination among members of 8 Vaccine Safety Datalink sites during September 1, 2022–March 31, 2023.

| Risk interval | Analyses | Relative incidence for ≥12 years old |  |  | Relative incidence for 12-64 years old |  |  | Relative incidence for ≥65 years old |  |  |
| --- | --- | --- | --- | --- | --- | --- | --- | --- | --- | --- |
|  |  | After COVID-19 vaccination | After influenza vaccination | After same-day coadministration | After COVID-19 vaccination | After influenza vaccination | After same-day coadministration | After COVID-19 vaccination | After influenza vaccination | After same-day coadministration |
| 1-21 days | Overall | 0.97 (0.66–1.42) | 0.94 (0.82–1.08) | 1.21 (0.95–1.55) | 1.00 (0.50–1.99) | 1.10 (0.87–1.40) | 0.97 (0.57–1.63) | 0.97 (0.61–1.53) | 0.89 (0.76–1.06) | 1.32 (1.00–1.74) |
|  | With prior SARS-CoV-2 infection<sup>1</sup> | 0.99 (0.34–2.94) | 1.04 (0.75–1.46) | 1.51 (0.83–2.75) | 3.05 (0.72–12.91) | 1.53 (0.96–2.42) | 1.43 (0.52–3.91) | 0.33 (0.04–2.53) | 0.77 (0.47–1.25) | 1.52 (0.73–3.20) |
|  | Without prior SARS-CoV-2 infection | 0.97 (0.65–1.47) | 0.92 (0.79–1.08) | 1.17 (0.89–1.53) | 0.75 (0.32–1.73) | 1.00 (0.75–1.32) | 0.87 (0.47–1.59) | 1.07 (0.67–1.72) | 0.91 (0.76–1.10) | 1.29 (0.95–1.74) |
| 1-42 days | Overall | 0.87 (0.61–1.23) | 0.93 (0.83–1.05) | 1.11 (0.91–1.36) | 0.89 (0.49–1.64) | 1.14 (0.93–1.39) | 1.02 (0.68–1.53) | 0.86 (0.56–1.32) | 0.87 (0.75–1.00) | 1.15 (0.91–1.46) |
|  | With prior SARS-CoV-2 infection<sup>1</sup> | 1.70 (0.73–3.96) | 1.12 (0.84–1.49) | 1.63 (0.97–2.73) | 3.28 (0.81–13.30) | 1.36 (0.89–2.09) | 2.09 (0.85–5.14) | 1.21 (0.41–3.62) | 0.98 (0.66–1.44) | 1.43 (0.76–2.71) |
|  | Without prior SARS-CoV-2 infection | 0.77 (0.52–1.15) | 0.90 (0.79–1.02) | 1.04 (0.84–1.30) | 0.68 (0.33–1.40) | 1.07 (0.85–1.36) | 0.86 (0.54–1.36) | 0.81 (0.51–1.30) | 0.85 (0.73–0.99) | 1.12 (0.87–1.44) |

<sup>1</sup> Had SARS-CoV-2 infection (i.e., SARS-CoV-2 positive laboratory test or a COVID-19 diagnosis) during the year prior (September 1, 2021–March 31, 2022).

Table 5 presents the results for Moderna bivalent vaccination and influenza vaccination, where no elevated risk was detected. Again, an elevated point estimate was observed but was not statistically significant and the confidence interval was wide, within 42 days after same-day coadministration (RI=3.08 [95% CI, 0.58–16.38]) in individuals aged 12-64 years with prior SARS-CoV-2 infection.

**Table 5.** Relative incidences of electronically identified first-in-1-year ischemic stroke in the 21 days and 42 days after Moderna bivalent COVID-19 vaccination and influenza vaccination among members of 8 Vaccine Safety Datalink sites during September 1, 2022–March 31, 2023.

| Risk interval | Analyses | Relative incidence for ≥12 years old |  |  | Relative incidence for 12-64 years old |  |  | Relative incidence for ≥65 years old |  |  |
| --- | --- | --- | --- | --- | --- | --- | --- | --- | --- | --- |
|  |  | After COVID-19 vaccination | After influenza vaccination | After same-day coadministration | After COVID-19 vaccination | After influenza vaccination | After same-day coadministration | After COVID-19 vaccination | After influenza vaccination | After same-day coadministration |
| 1-21 days | Overall | 0.65 (0.35–1.23) | 0.96 (0.83–1.11) | 0.87 (0.52–1.46) | 0.40 (0.13–1.25) | 1.02 (0.79–1.32) | 0.69 (0.28–1.68) | 0.81 (0.38–1.73) | 0.95 (0.80–1.12) | 0.97 (0.52–1.82) |
|  | With prior SARS-CoV-2 infection<sup>1</sup> | NA | 1.04 (0.73–1.49) | 0.71 (0.18–2.78) | NA | 1.23 (0.72–2.09) | 0.78 (0.14–4.31) | NA | 0.93 (0.58–1.50) | 0.66 (0.08–5.37) |
|  | Without prior SARS-CoV-2 infection | 0.79 (0.42–1.50) | 0.94 (0.81–1.10) | 0.90 (0.52–1.56) | 0.54 (0.17–1.78) | 0.97 (0.72–1.30) | 0.66 (0.24–1.84) | 0.92 (0.43–1.96) | 0.95 (0.79–1.14) | 1.02 (0.53–1.97) |
| 1-42 days | Overall | 0.95 (0.59–1.52) | 0.97 (0.86–1.10) | 1.12 (0.77–1.63) | 1.18 (0.55–2.51) | 1.14 (0.92–1.40) | 1.30 (0.68–2.47) | 0.82 (0.44–1.51) | 0.91 (0.79–1.05) | 1.05 (0.66–1.68) |
|  | With prior SARS-CoV-2 infection<sup>1</sup> | 0.56 (0.16–2.01) | 1.17 (0.87–1.58) | 1.26 (0.42–3.78) | 0.64 (0.11–3.78) | 1.36 (0.86–2.16) | 3.08 (0.58–16.38) | 0.41 (0.09–2.00) | 1.05 (0.71–1.54) | 0.60 (0.12–3.11) |
|  | Without prior SARS-CoV-2 infection | 1.03 (0.62–1.72) | 0.94 (0.83–1.07) | 1.10 (0.74–1.65) | 1.46 (0.63–3.36) | 1.09 (0.86–1.38) | 1.08 (0.53–2.20) | 0.86 (0.45–1.65) | 0.89 (0.76–1.04) | 1.12 (0.69–1.83) |

<sup>1</sup> Had SARS-CoV-2 infection (i.e., SARS-CoV-2 positive laboratory test or a COVID-19 diagnosis) during the year prior (September 1, 2021–March 31, 2022). NA, not available due to extremely low case count.

### 3.4. Exploratory outcomes first-ever and first-in-1-year ischemic stroke or TIA

We did not observe an elevated risk associated with either vaccination alone or same-day coadministration. This finding was consistent across vaccine products and in both the overall and subgroup analyses (Tables S3-S6).

Because no elevated risk was detected in the analyses of electronically identified study outcomes (i.e., the lower bound of the 95% CI for the RI did not exceed 1.00), no chart reviews or analyses restricted to chart-confirmed outcomes were conducted.

## 4. DISCUSSION

In this U.S. multi-site SCCS study, we assessed the risk of ischemic stroke following COVID-19 bivalent vaccination, influenza vaccination, and their coadministration on the same day. Our findings differ from the original statistical signal detected by VSD’s concurrent-comparator RCA, which observed an elevated adjusted rate ratio early in the season for ischemic events (i.e., ischemic stroke or TIA) following the Pfizer-BioNTech COVID-19 bivalent booster among adults aged 65 years and older. By the end of the season, however, the RCA estimates had attenuated and were closer to the null, aligning more closely with the results of this study. The difference in findings is expected given that the RCA method compares vaccinated individuals with concurrent comparators who were vaccinated earlier and can be affected by residual confounding, whereas the SCCS design compares individuals to themselves over time and implicitly adjusts for person-specific time-invariant characteristics. We found no association between first-ever ischemic stroke and either Pfizer-BioNTech or Moderna COVID-19 bivalent vaccinations, influenza vaccination, or their same-day coadministration with risk intervals of 1-21 and 1-42 days in the overall analyses and subgroup analyses by age (12-64 and ≥65 years) and history of SARS-CoV-2 infection. For the secondary and exploratory outcomes—first-in-1-year ischemic stroke, and first-ever and first-in-1-year ischemic stroke or TIA, respectively—no elevated risk was observed following either vaccination alone or same-day coadministration, and this was consistent across bivalent vaccine type and subgroups.

A small number of stratified analyses showed RIs below 1.00 with confidence intervals excluding 1.00, including influenza vaccination in the 1–42-day risk interval among adults aged ≥65 years without prior SARS-CoV-2 infection (RI = 0.83; 95% CI, 0.70–0.97; Table 2) and Moderna bivalent COVID-19 vaccination in the 1–21-day risk interval in the overall analysis (RI = 0.45; 95% CI, 0.20–0.98; Table 3). These findings should be interpreted cautiously, as they may reflect residual confounding, healthy-vaccinee effects, or chance due to multiple comparisons rather than a true protective effect.

We focused on ischemic stroke in this study, because unlike TIA (which are by definition transient), ischemic stroke poses a more severe and lasting health risk [25]. Additionally, the diagnosis of ischemic stroke carries greater certainty than TIA, as it is more likely to be supported by objective evidence such as imaging findings. Because previous studies have shown that the risk of ischemic events (ischemic stroke or TIA) is significantly associated with COVID-19 disease itself [19,20], it was important to assess whether COVID-19 vaccination was also associated with the risk of more severe outcomes like ischemic stroke.

Our results are consistent with most earlier studies in showing no increased risk of ischemic events after bivalent COVID-19 vaccination alone (without same-day coadministration of influenza vaccine, but allowing other vaccines), influenza vaccination alone (without same-day coadministration of bivalent COVID-19 vaccine, but allowing other vaccines), or their same-day coadministration. The higher risk reported by Lu et al. [17] pertains specifically to coadministration involving high-dose or adjuvanted influenza vaccines, while our analysis included any influenza vaccine; therefore, their findings are not directly comparable to ours. Unlike earlier studies that examined only COVID-19 vaccines [9,10,15,16], our SCCS analyses simultaneously assessed risks for bivalent vaccination alone, influenza vaccination alone, and same-day coadministration. In addition, all but Xu et al. [18] did not account for prior SARS-CoV-2 infection when evaluating stroke risk. Our findings are also consistent with a large nationwide cohort study from Denmark that evaluated the safety of bivalent mRNA COVID-19 booster vaccines among adults aged ≥50 years using registry-based data [26]. In that study, bivalent vaccination was not associated with an increased risk of ischemic stroke or other serious adverse events in the 28 days following vaccination, consistent with our analyses.

A recent single-site, method-focused SCCS study reported a statistically significant association between Pfizer-BioNTech bivalent COVID-19 vaccination and ischemic event during periods of overlapping exposure among individuals with a prior-year history of SARS-CoV-2 infection, based on ICD-10–identified outcomes [27]. That study was designed primarily to explore methodological considerations and potential effect modification, and the observed association was limited to a narrowly defined subgroup with wide confidence intervals, indicating statistical instability. In contrast, the present multi-site VSD study included substantially larger numbers of events across eight health systems and did not identify a statistically significant increased risk following same-day coadministration, including among individuals with prior SARS-CoV-2 infection. Consequently, no further characterization of risk after same-day coadministration was warranted.

The current study has limitations. First, while we employed the event-dependent SCCS method to account for the impact of a prior outcome event on the probability of future vaccination, this method could not accommodate cases of vaccine coadministration that did not occur on the same-day but occurred in close proximity with overlapping risk intervals. However, given that same-day coadministration was not statistically significantly associated with an increased risk of study outcomes, it is biologically plausible that vaccine coadministration in close-proximity would also not be associated with an increased risk. Second, ischemic events were identified through ICD-10-CM codes without medical chart confirmation, and thus misclassification of ischemic events and symptom onset dates was possible. Such outcome misclassification, as noted in previous vaccine safety studies, could bias our risk estimates [28,29]. Third, although we excluded ischemic events that occurred within 30 days of SARS-CoV-2 infection due to the elevated risk of ischemic events after SARS-CoV-2 infection [19,20,30] and conducted subgroup analysis based on history of SARS-CoV-2 infection in the year prior to September 1, 2022, there was a possibility of misclassification due to incomplete information on SARS-CoV-2 infection.Results of home-administered rapid tests were not captured, and some individuals may not have tested or sought care. Fourth, we did not exclude ischemic strokes potentially related to influenza-like illness [31], which could have influenced the results. Additionally, misclassification of vaccination status was possible for the bivalent COVID-19 vaccines and the influenza vaccines, particularly if vaccinations were received at facilities outside these VSD sites and were not captured in linked immunization information systems. However, such misclassification was expected to be minimal because VSD sites use comprehensive electronic health record systems, and external vaccinations administered in community pharmacies or other healthcare settings were routinely captured through state immunization information systems and incorporated into the VSD’s standardized data model [32]. Lastly, some subgroup analyses yielded wide confidence intervals due to small numbers of events after stratification by age, prior SARS-CoV-2 infection, and coadministration with influenza vaccination, limiting precision and warranting cautious interpretation of elevated point estimates.

In conclusion, in this multi-site study, we did not find evidence of an increased risk of ischemic stroke in the overall analyses or in subgroup analyses by age and history of SARS-CoV-2 infection following bivalent mRNA COVID-19 vaccination, influenza vaccination, or their coadministration. These findings align with previous studies that have not observed a significant association between bivalent COVID-19 vaccination and ischemic stroke. The safety of updated COVID-19 vaccine formulations should continue to be monitored.

## Data availability

The data used in this study are not publicly available due to privacy restrictions but may be available from the corresponding author on reasonable request, subject to institutional approvals.

---

**Table S1.** Case counts of ischemic stroke and transient ischemic attack and inclusion in SCCS analyses for 1–21-day and 1–42-day risk intervals among recipients of Pfizer-BioNTech bivalent COVID-19 and influenza vaccines at 8 Vaccine Safety Datalink sites during September 1, 2022–March 31, 2023.

| Outcomes | Total vaccinated cases <sup>1</sup> | Day 0 cases <sup>2</sup> | 1–21-day risk interval |  | 1–42-day risk interval |  |
| --- | --- | --- | --- | --- | --- | --- |
|  |  |  | Cases with coadministration within 1–21 days apart <sup>3</sup> | Cases included in SCCS analyses <sup>4</sup> | Cases with coadministration within 1–42 days apart <sup>3</sup> | Cases included in SCCS analyses <sup>4</sup> |
| First-ever ischemic stroke | 4,009 | 89 | 452 | 3,468 | 751 | 3,169 |
| First-in-1-year ischemic stroke | 4,435 | 97 | 491 | 3,847 | 819 | 3,519 |
| First-ever ischemic stroke or transient ischemic attack | 5,098 | 118 | 622 | 4,358 | 1,004 | 3,976 |
| First-in-1-year ischemic stroke or transient ischemic attack | 5,889 | 139 | 709 | 5,041 | 1,147 | 4,603 |

<sup>1</sup> Cases who received Pfizer-BioNTech bivalent COVID-19 vaccine alone, or influenza vaccine alone, or both during observation period; <sup>2</sup> cases that occurred on dates of either COVID-19 vaccination or influenza vaccination; <sup>3</sup> cases who received Pfizer-BioNTech bivalent COVID-19 vaccine and influenza vaccine within 1–21 or 1–42 days apart, not including same-day coadministration; <sup>4</sup> cases in SCCS analyses including recipients of Pfizer-BioNTech bivalent COVID-19 vaccine alone, influenza vaccine alone, same-day coadministration, or who received Pfizer-BioNTech bivalent COVID-19 vaccine and influenza vaccine ≥22 or ≥43 days apart.

**Table S2.** Case counts of ischemic stroke and transient ischemic attack and inclusion in SCCS analyses for 1–21-day and 1–42-day risk intervals among recipients of Moderna bivalent COVID-19 and influenza vaccines at 8 Vaccine Safety Datalink sites during September 1, 2022–March 31, 2023.

| Outcomes | Total vaccinated cases <sup>1</sup> | Day 0 cases <sup>2</sup> | 1–21-day risk interval |  | 1–42-day risk interval |  |
| --- | --- | --- | --- | --- | --- | --- |
|  |  |  | Cases with coadministration within 1–21 days apart <sup>3</sup> | Cases included in SCCS analyses <sup>4</sup> | Cases with coadministration within 1–42 days apart <sup>3</sup> | Cases included in SCCS analyses <sup>4</sup> |
| First-ever ischemic stroke | 3,064 | 81 | 252 | 2,731 | 414 | 2,569 |
| First-in-1-year ischemic stroke | 3,406 | 88 | 276 | 3,042 | 455 | 2,863 |
| First-ever ischemic stroke or transient ischemic attack | 3,863 | 107 | 333 | 3,423 | 545 | 3,211 |
| First-in-1-year ischemic stroke or transient ischemic attack | 4,472 | 124 | 384 | 3,964 | 628 | 3,720 |

<sup>1</sup> Cases who received Moderna bivalent COVID-19 vaccine alone, or influenza vaccine alone, or both during observation period; <sup>2</sup> cases that occurred on dates of either COVID-19 vaccination or influenza vaccination; <sup>3</sup> cases who received Moderna bivalent COVID-19 vaccine and influenza vaccine within 1–21 or 1–42 days apart, not including same-day coadministration; <sup>4</sup> cases in SCCS analyses including recipients of Moderna bivalent COVID-19 vaccine alone, influenza vaccine alone, same-day coadministration, or who received Moderna bivalent COVID-19 vaccine and influenza vaccine ≥22 or ≥43 days apart.

**Table S3.** Relative incidences of electronically identified first-ever ischemic stroke or transient ischemic attack in the 21 days and 42 days after Pfizer-BioNTech bivalent COVID-19 vaccination and influenza vaccination among members of 8 Vaccine Safety Datalink sites during September 1, 2022–March 31, 2023.

|  |  | Relative incidence for ≥12 years old |  |  | Relative incidence for 12-64 years old |  |  | Relative incidence for ≥65 years old |  |  |
| --- | --- | --- | --- | --- | --- | --- | --- | --- | --- | --- |
| Risk interval | Analyses | After COVID-19 vaccination | After influenza vaccination | After same-day coadministration | After COVID-19 vaccination | After influenza vaccination | After same-day coadministration | After COVID-19 vaccination | After influenza vaccination | After same-day coadministration |
| 1-21 days | Overall | 1.12 (0.80–1.56) | 0.96 (0.84–1.09) | 1.09 (0.86–1.38) | 1.01 (0.56–1.83) | 1.01 (0.80–1.26) | 0.88 (0.53–1.43) | 1.18 (0.78–1.77) | 0.96 (0.82–1.12) | 1.19 (0.91–1.56) |
|  | With prior SARS-CoV-2 infection <sup>1</sup> | 1.28 (0.55–2.97) | 1.03 (0.76–1.40) | 1.29 (0.70–2.37) | 2.17 (0.75–6.24) | 1.32 (0.85–2.05) | 1.18 (0.42–3.31) | 0.63 (0.14–2.84) | 0.88 (0.57–1.35) | 1.36 (0.64–2.91) |
|  | Without prior SARS-CoV-2 infection | 1.10 (0.76–1.58) | 0.94 (0.82–1.09) | 1.07 (0.83–1.37) | 0.77 (0.37–1.61) | 0.93 (0.71–1.21) | 0.82 (0.47–1.43) | 1.27 (0.83–1.94) | 0.97 (0.82–1.15) | 1.17 (0.88–1.56) |
| 1-42 days | Overall | 0.91 (0.65–1.26) | 0.94 (0.84–1.05) | 0.90 (0.74–1.09) | 0.84 (0.49–1.43) | 1.07 (0.89–1.30) | 0.90 (0.62–1.30) | 0.95 (0.63–1.43) | 0.90 (0.79–1.03) | 0.91 (0.72–1.15) |
|  | With prior SARS-CoV-2 infection <sup>1</sup> | 1.40 (0.68–2.90) | 1.11 (0.85–1.44) | 1.14 (0.68–1.92) | 1.18 (0.40–3.46) | 1.13 (0.75–1.69) | 1.52 (0.66–3.46) | 1.64 (0.62–4.35) | 1.13 (0.80–1.60) | 0.98 (0.49–1.93) |
|  | Without prior SARS-CoV-2 infection | 0.82 (0.57–1.19) | 0.91 (0.81–1.03) | 0.87 (0.70–1.08) | 0.76 (0.41–1.43) | 1.06 (0.85–1.31) | 0.80 (0.52–1.22) | 0.85 (0.54–1.35) | 0.87 (0.75–1.00) | 0.90 (0.71–1.15) |

<sup>1</sup> Had SARS-CoV-2 infection (i.e., SARS-CoV-2 positive laboratory test or a COVID-19 diagnosis) during the year prior (September 1, 2021–March 31, 2022).

**Table S4.**
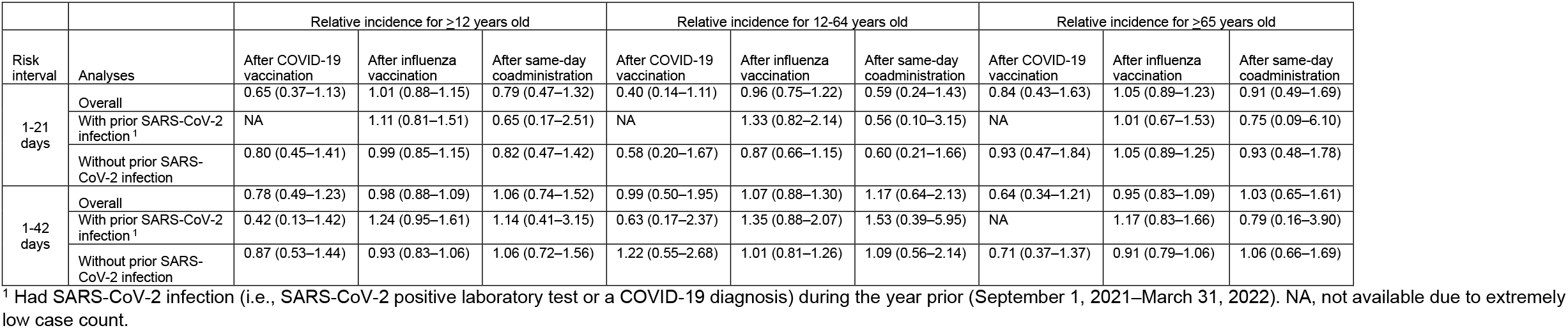
Relative incidences of electronically identified first-ever ischemic stroke or transient ischemic attack in the 21 days and 42 days after Moderna bivalent COVID-19 vaccination and influenza vaccination among members of 8 Vaccine Safety Datalink sites during September 1, 2022–March 31, 2023.

**Table S5.** Relative incidences of electronically identified first-in-1-year ischemic stroke or transient ischemic attack in the 21 days and 42 days after Pfizer-BioNTech bivalent COVID-19 vaccination and influenza vaccination among members of 8 Vaccine Safety Datalink sites during September 1, 2022–March 31, 2023.

|  |  | Relative incidence for ≥12 years old |  |  | Relative incidence for 12-64 years old |  |  | Relative incidence for ≥65 years old |  |  |
| --- | --- | --- | --- | --- | --- | --- | --- | --- | --- | --- |
| Risk interval | Analyses | After COVID-19 vaccination | After influenza vaccination | After same-day coadministration | After COVID-19 vaccination | After influenza vaccination | After same-day coadministration | After COVID-19 vaccination | After influenza vaccination | After same-day coadministration |
| 1-21 days | Overall | 1.11 (0.81–1.52) | 0.99 (0.88–1.11) | 1.13 (0.91–1.40) | 0.93 (0.52–1.67) | 1.01 (0.81–1.25) | 0.97 (0.61–1.53) | 1.21 (0.83–1.76) | 1.00 (0.86–1.15) | 1.21 (0.94–1.54) |
|  | With prior SARS-CoV-2 infection <sup>1</sup> | 1.09 (0.48–2.49) | 0.96 (0.72–1.28) | 1.46 (0.86–2.50) | 1.87 (0.67–5.20) | 1.28 (0.84–1.94) | 1.59 (0.67–3.75) | 0.53 (0.12–2.33) | 0.78 (0.52–1.18) | 1.40 (0.70–2.78) |
|  | Without prior SARS-CoV-2 infection | 1.12 (0.80–1.57) | 0.99 (0.87–1.13) | 1.09 (0.86–1.37) | 0.71 (0.34–1.49) | 0.94 (0.73–1.20) | 0.84 (0.49–1.44) | 1.32 (0.89–1.94) | 1.03 (0.88–1.20) | 1.18 (0.91–1.54) |
| 1-42 days | Overall | 0.94 (0.70–1.27) | 0.96 (0.87–1.06) | 0.97 (0.81–1.17) | 0.83 (0.49–1.40) | 1.07 (0.89–1.28) | 0.94 (0.66–1.34) | 1.01 (0.69–1.46) | 0.93 (0.82–1.05) | 1.00 (0.81–1.23) |
|  | With prior SARS-CoV-2 infection <sup>1</sup> | 1.33 (0.67–2.64) | 1.07 (0.84–1.37) | 1.42 (0.90–2.25) | 1.02 (0.36–2.89) | 1.10 (0.75–1.62) | 1.89 (0.88–4.03) | 1.65 (0.67–4.06) | 1.07 (0.78–1.48) | 1.24 (0.68–2.24) |
|  | Without prior SARS-CoV-2 infection | 0.88 (0.63–1.23) | 0.93 (0.84–1.05) | 0.92 (0.76–1.12) | 0.79 (0.43–1.45) | 1.06 (0.86–1.30) | 0.78 (0.52–1.19) | 0.92 (0.61–1.39) | 0.90 (0.79–1.03) | 0.97 (0.78–1.22) |

<sup>1</sup> Had SARS-CoV-2 infection (i.e., SARS-CoV-2 positive laboratory test or a COVID-19 diagnosis) during the year prior (September 1, 2021–March 31, 2022).

**Table S6.** Relative incidences of electronically identified first-in-1-year ischemic stroke or transient ischemic attack in the 21 days and 42 days after Moderna bivalent COVID-19 vaccination and influenza vaccination among members of 8 Vaccine Safety Datalink sites during September 1, 2022–March 31, 2023.

|  |  | Relative incidence for ≥12 years old |  |  | Relative incidence for 12-64 years old |  |  | Relative incidence for ≥65 years old |  |  |
| --- | --- | --- | --- | --- | --- | --- | --- | --- | --- | --- |
| Risk interval | Analyses | After COVID-19 vaccination | After influenza vaccination | After same-day coadministration | After COVID-19 vaccination | After influenza vaccination | After same-day coadministration | After COVID-19 vaccination | After influenza vaccination | After same-day coadministration |
| 1-21 days | Overall | 0.79 (0.47–1.31) | 1.02 (0.90–1.15) | 0.85 (0.53–1.35) | 0.38 (0.14–1.06) | 0.98 (0.78–1.23) | 0.68 (0.30–1.56) | 1.06 (0.59–1.90) | 1.05 (0.91–1.22) | 0.94 (0.54–1.65) |
|  | With prior SARS-CoV-2 infection <sup>1</sup> | NA | 1.03 (0.77–1.39) | 0.75 (0.24–2.38) | NA | 1.28 (0.81–2.02) | 1.21 (0.29–5.00) | NA | 0.92 (0.62–1.36) | 0.45 (0.06–3.46) |
|  | Without prior SARS-CoV-2 infection | 0.97 (0.58–1.62) | 1.02 (0.89–1.16) | 0.87 (0.52–1.44) | 0.54 (0.19–1.55) | 0.90 (0.69–1.18) | 0.55 (0.20–1.51) | 1.20 (0.67–2.17) | 1.08 (0.92–1.26) | 1.02 (0.57–1.83) |
| 1-42 days | Overall | 0.83 (0.54–1.27) | 1.00 (0.90–1.11) | 1.04 (0.74–1.45) | 0.92 (0.47–1.82) | 1.09 (0.90–1.31) | 1.26 (0.71–2.23) | 0.76 (0.44–1.34) | 0.97 (0.86–1.10) | 0.97 (0.64–1.47) |
|  | With prior SARS-CoV-2 infection <sup>1</sup> | 0.52 (0.19–1.46) | 1.16 (0.90–1.48) | 0.92 (0.36–2.32) | 0.65 (0.17–2.44) | 1.29 (0.86–1.93) | 2.08 (0.58–7.44) | 0.35 (0.07–1.67) | 1.10 (0.80–1.52) | 0.41 (0.09–1.93) |
|  | Without prior SARS-CoV-2 infection | 0.90 (0.57–1.44) | 0.97 (0.87–1.09) | 1.06 (0.74–1.52) | 1.09 (0.49–2.38) | 1.04 (0.84–1.28) | 1.10 (0.58–2.09) | 0.82 (0.45–1.47) | 0.95 (0.83–1.09) | 1.06 (0.69–1.64) |

<sup>1</sup> Had SARS-CoV-2 infection (i.e., SARS-CoV-2 positive laboratory test or a COVID-19 diagnosis) during the year prior (September 1, 2021–March 31, 2022). NA, not available due to extremely low case count.

